# Unpublished clinical studies in pediatric emergence delirium – a cross sectional analysis

**DOI:** 10.1101/2020.01.24.20018663

**Authors:** Jochen Meyburg, Markus Ries

## Abstract

**Objectives:** Emergence delirium (ED) is a frequent and potentially serious complication of general anesthesia in children. Although there are various treatment strategies, no general management recommendations can be made. Selective reporting of study results may impair clinical decision making. We therefore analyzed whether the results of completed registered clinical studies in patients with pediatric ED are publically available or remain unpublished.

**Design:** Cross sectional analysis.

**Setting:** ClinicalTrials.gov and ClinicalTrialsRegister.eu

**Participants and outcome measures:** We determined the proportion of published and unpublished studies registered at ClinicalTrials.gov and ClinicalTrialsRegister.eu that were marked as completed by September 1^st^ 2018. The major trial and literature databases were used to search for publications. In addition, the study investigators were contacted directly.

**Results:** Of the 44 registered studies on pediatric ED, only 24 (54%) have been published by September 2019. Published trials contained data from n=2556 patients, whereas n=1644 patients were enrolled in unpublished trials. Median time to publication was 19 months. Studies completed in recent years were published faster, but still only 9 of 25 trials were published within 12 months after completion.

**Conclusion:** There is a distinct publication gap in clinical research in pediatric ED that may have an impact on meta-analyses and clinical practice.

**Strengths and limitations of this study:**

- This study quantitates the amount of research waste in pediatric emergence delirium assessed as a) the number and b) sample sizes of published and unpublished completed clinical studies
- The precise reasons for non-publication of the studies included in this analysis remain unknown
- Strengths of findings as well as directions of individual unpublished studies remain unknown
- Study registers other than ClinicalTrials.gov and ClinicalTrialsRegister.eu were not analyzed

**Funding statement:** This research received no specific grant from any funding agency in the public, commercial or not-for-profit sectors.

## Introduction

Emergence delirium (ED) can be a very stressful event for both patients and caregivers during general anesthesia in children. Although it may also develop in adults, ED is much more common in pediatric patients, with prevalences between 25% and 80% depending on the definition of ED ^1^. Symptoms usually begin shortly after emergence from anesthesia and can be very frightening including self-inflicted injury or accidental removal of catheters and other medical devices. Although episodes of ED are usually short lived, it has been suspected that ED may be associated with long-term behavioral disturbances such as eating disorders, sleeping disorders, and separation anxiety^2^.

The exact pathophysiology of ED is not yet understood. However, several risk factors are known: young age, use of volatile anesthetics (especially sevoflurane), type of surgery (increased risk for otorhinolaryngeal and ophthalmological procedures), parental as well as patient anxiety, and pre-existing behavioral problems ^3^. Whereas anxiety and behavioral problems can be addressed by non-pharmacological interventions, most of these risk factors cannot be modified and prompt the pre- and/or perioperative administration of various medications including benzodiazepines, alpha-2-agonists, propofol, opioids, and ketamine ^4 5^.

However, although it is evident that all of these drugs may have beneficial effects in specific settings to reduce the rates of ED, no universal recommendations can be derived from the existing literature for this very common and potentially serious complication. This is a typical situation in the treatment of pediatric patients, where many treatment decisions are still based on incomplete clinical data, and off-label use of various drugs is common. One important factor for the lack of clinical consensus data might be a publication bias. It is twice as likely that a positive outcome of an intervention is reported than a negative one ^6^. Such selective reporting of positive results is likely to influence clinical decision making. We therefore investigated potential publication bias and time to publication in registered clinical trials on ED in children.

## Methods

### Identification of clinical trials

Two databases were assessed to identify registered clinical trials on Pediatric Emergence Delirium reported as completed: 1) the ClinicalTrials.gov database provided by the U.S. National Library of Medicine and 2) the European Union Clinical Trials Register at ClinicalTrialsRegister.eu. Search criteria were: keywords “emergence delirium” and “emergence agitation” with the query selection parameters “completed studies” and “child (0-17 years)”. Close of database was September 1 ^st^ 2019. Data were downloaded for further analysis.

### Search for publications of completed trials

To identify publications related to the registered and completed trials, ClinicalTrials.gov, PubMed and Google Scholar were searched for NCT number, EudraCT number, study title, principal investigator, study sponsor and keywords generated from the study title. If no respective publication was found, the principal investigators were contacted by email and/or ResearchGate and asked to provide information whether the study was published in a source not covered by PubMed or Google Scholar.

### Data Analysis

The STROBE criteria (STrengthening the Reporting of OBservational studies in Epidemiology) were applied for design and analysis of this study ^7^. Data were analyzed for age and number of participants, gender, study type, study design, condition, intervention, availability of study results, completion date, publication date, sponsor and country of sponsor. Trials were categorized into eight groups according to their main research topic. Time to publication was calculated as the difference in months between study completion date and publication date and. Missing data were not imputed. All statistical analyses were performed in SPSS 20 (IBM Corporations, Armonk, New York) using standard methods for descriptive statistics.

## Results

### Publication status of studies

We identified a total of 47 studies that were reported as completed in the two trial databases. Of these, three unpublished studies were completed less than one year before close of the database. Because the U.S. Food and Drug Administration (FDA) allows a time frame of one year between completion and publication of the study as specified in the in the FDA Drug Administration Amendments Act (FDAAA) ^8^, these three studies were excluded from the analysis. Of the remaining 44 studies, 29 were published and 19 were unpublished. Nine principal investigators of the unpublished studies could not be contacted by email or the ResearchGate social network. Of the remaining ten, two replied and confirmed that the study results had not been published yet (figure 1). Publication rates considerably varied between different countries of the sponsor (table 1) and main topics of the investigations (table 2).

**Table 1:** Published (n=25) and unpublished (n=19) completed studies on pediatric emergence delirium by country

| Countries | Published studies<br>(n) | Unpublished studies<br>(n) |
| --- | --- | --- |
| Belgium | 2 | 0 |
| Brasil | 0 | 1 |
| Canada | 1 | 0 |
| China | 0 | 2 |
| Egypt | 1 | 0 |
| Greece | 1 | 0 |
| India | 1 | 1 |
| Italy | 1 | 0 |
| Kenya | 0 | 1 |
| South Korea | 8 | 4 |
| Thailand | 2 | 1 |
| Turkey | 3 | 2 |
| United States | 4 | 8 |

**Table 2:** Publication status of studies registered as completed on ClinicalTrials.gov and ClinicalTrialsRegister.eu involving children with emergence delirium

| **Issue** | **Overall number of studies** | **Number and percentage of published studies** | **Number of patients enrolled in unpublished studies** |
| --- | --- | --- | --- |
| Dexmedetomidin | 13 | 5 (38%) | 598 |
| Diagnostic criteria | 6 | 2 (33%) | 326 |
| Non-pharmacological interventions | 5 | 4 (80%) | 100 |
| Opioids | 5 | 2 (40%) | 322 |
| Other drugs | 5 | 4 (80%) | 66 |
| Propofol | 4 | 3 (75%) | 100 |
| Volatile anesthetics | 3 | 1 (33%) | 132 |
| Midazolam | 3 | 3 (100%) | 0 |

**Figure 1:**
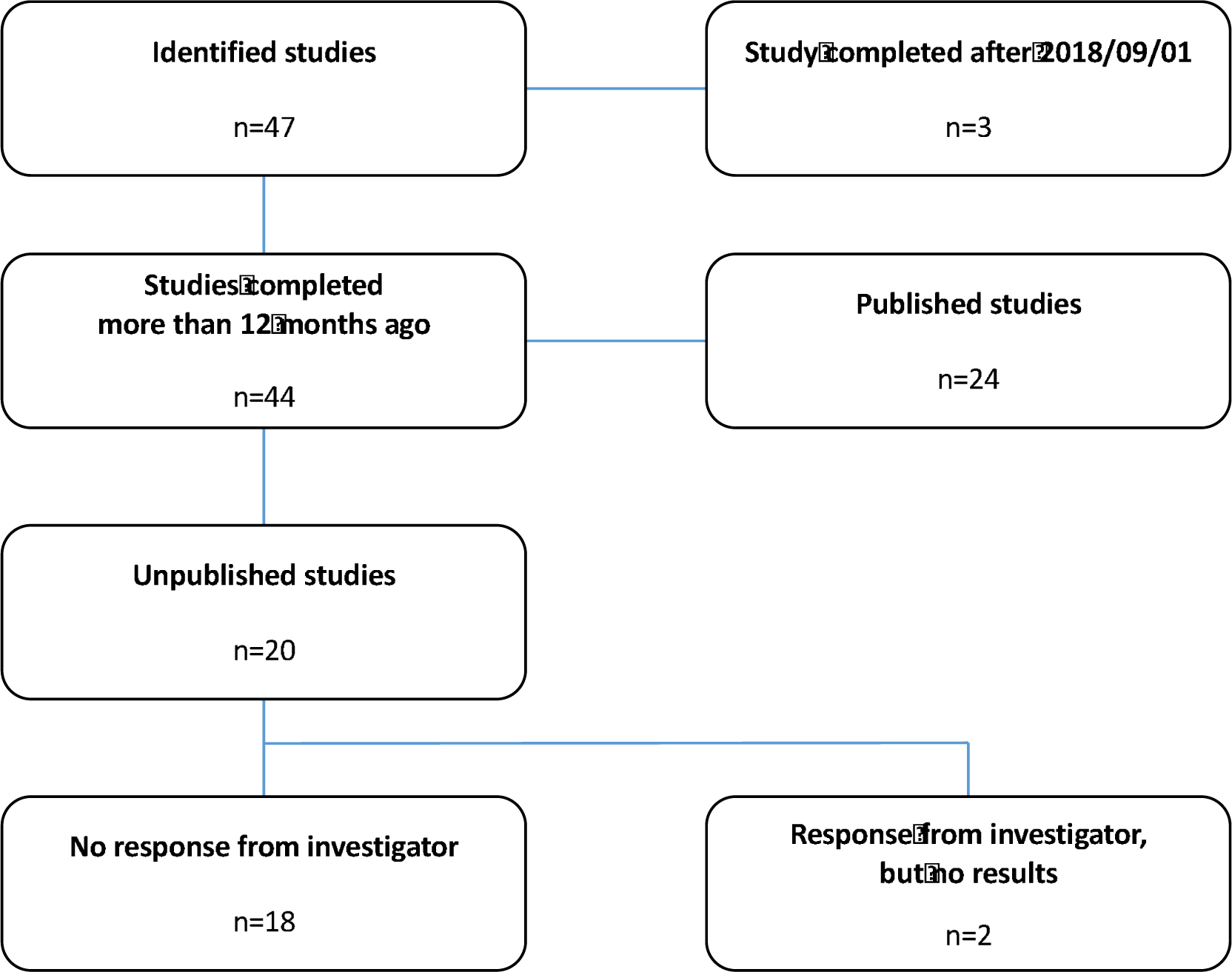
Flowsheet: details of the study selection process.

The numbers of published and unpublished studies for each year of study completion (2007 – 2018) is shown in figure 2. An increasing number of publications over the years can be observed as well as an increasing proportion of unpublished studies which even exceeded the number of published studies in the last three years.

**Figure 2:**
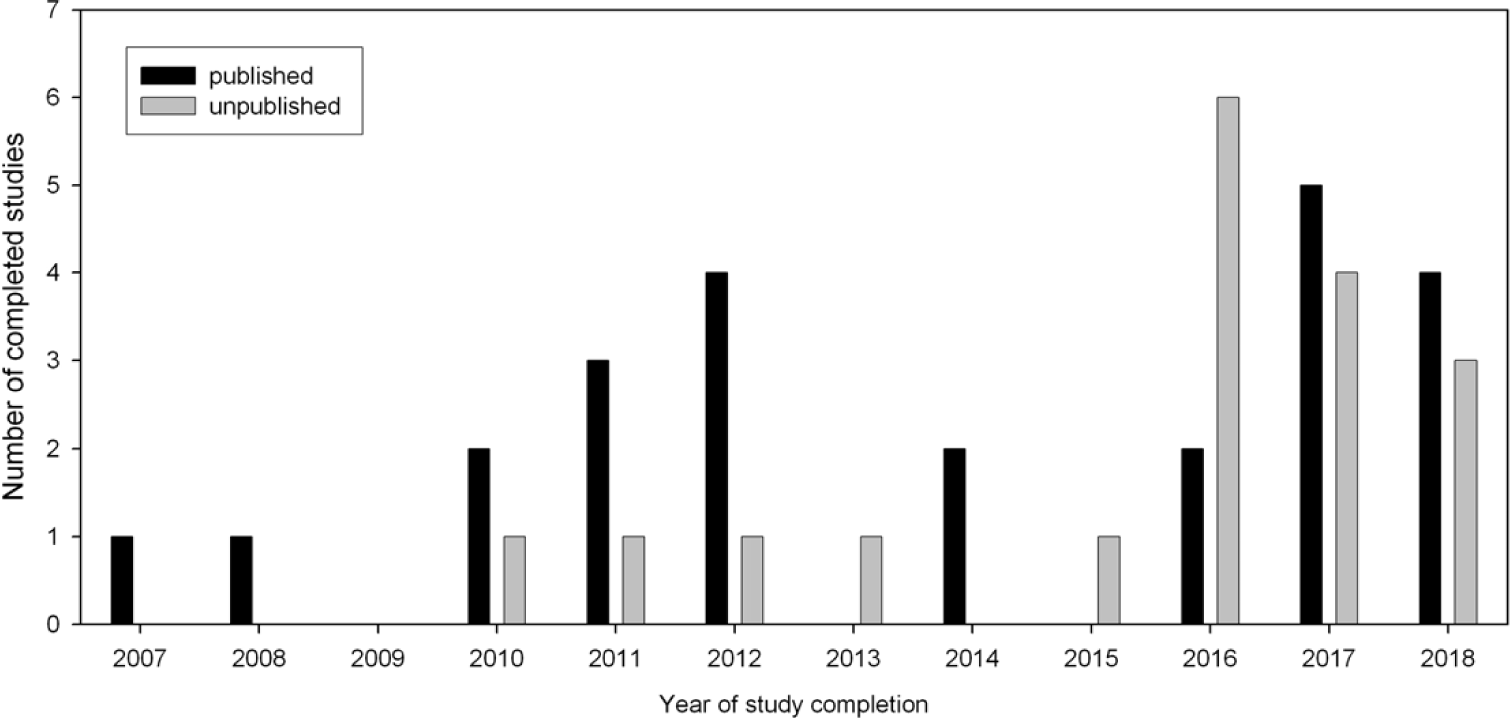
Distribution of published (n=24) and unpublished (n=20) trials by year of completion.

### Patient numbers

All studies involved both genders. Published trials contained data from n=2556 patients, whereas n=1644 patients were enrolled in unpublished trials. Median size of published trials was 90 (IQR 68-136), range 40-418, whereas median size of unpublished trials was 80 (IQR 55-100), range 22-156 participants. Of note, the number of patients enrolled in unpublished studies significantly exceeded those in published studies during the last years (figure 3).

**Figure 3:**
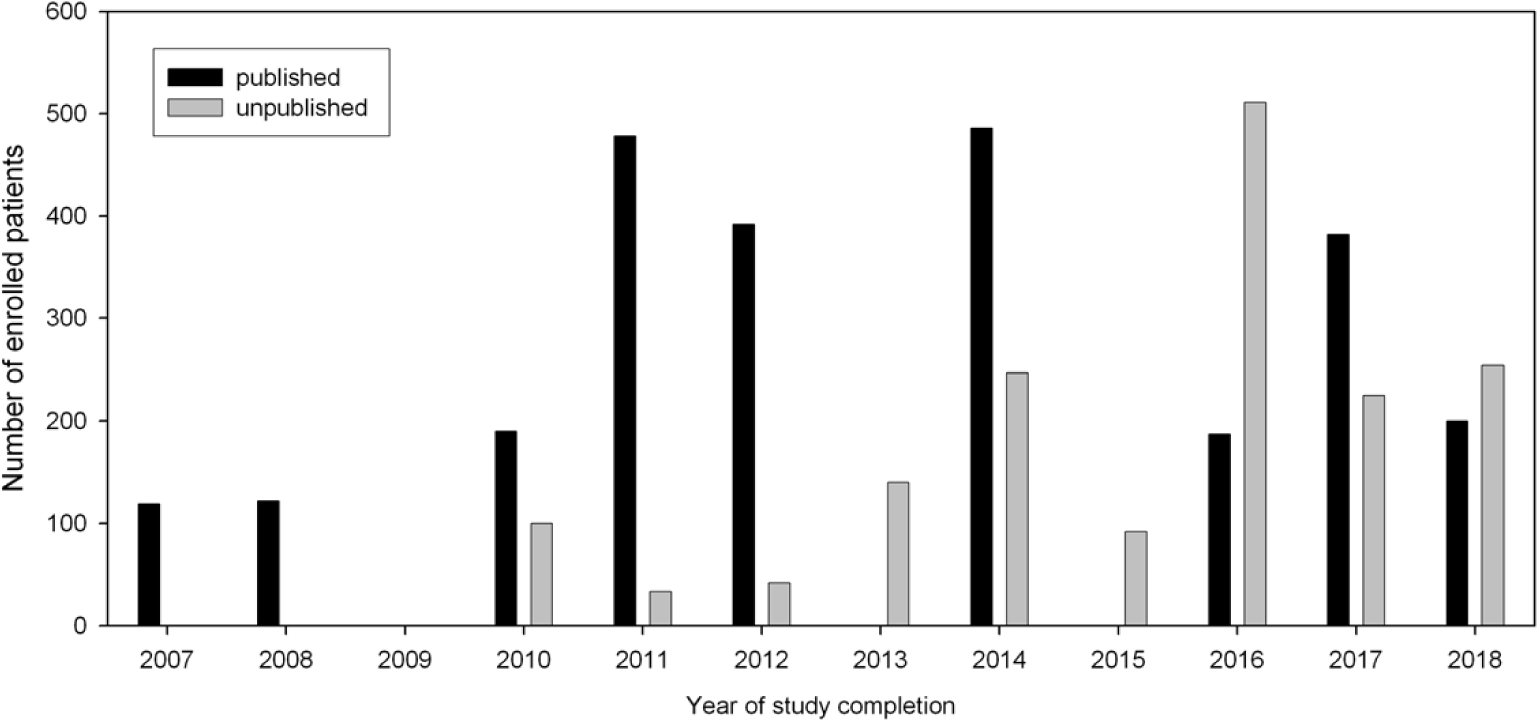
Distribution of patient count stratified by publication status and year.

### Time to publication

Median time to publication was 19 (IQR 12-27), range 3 to 104 months. More recent studies were published faster, but still only 9 of 25 trials were published within 12 months after completion as warranted by the FDAAA (figure 4).

**Figure 4:**
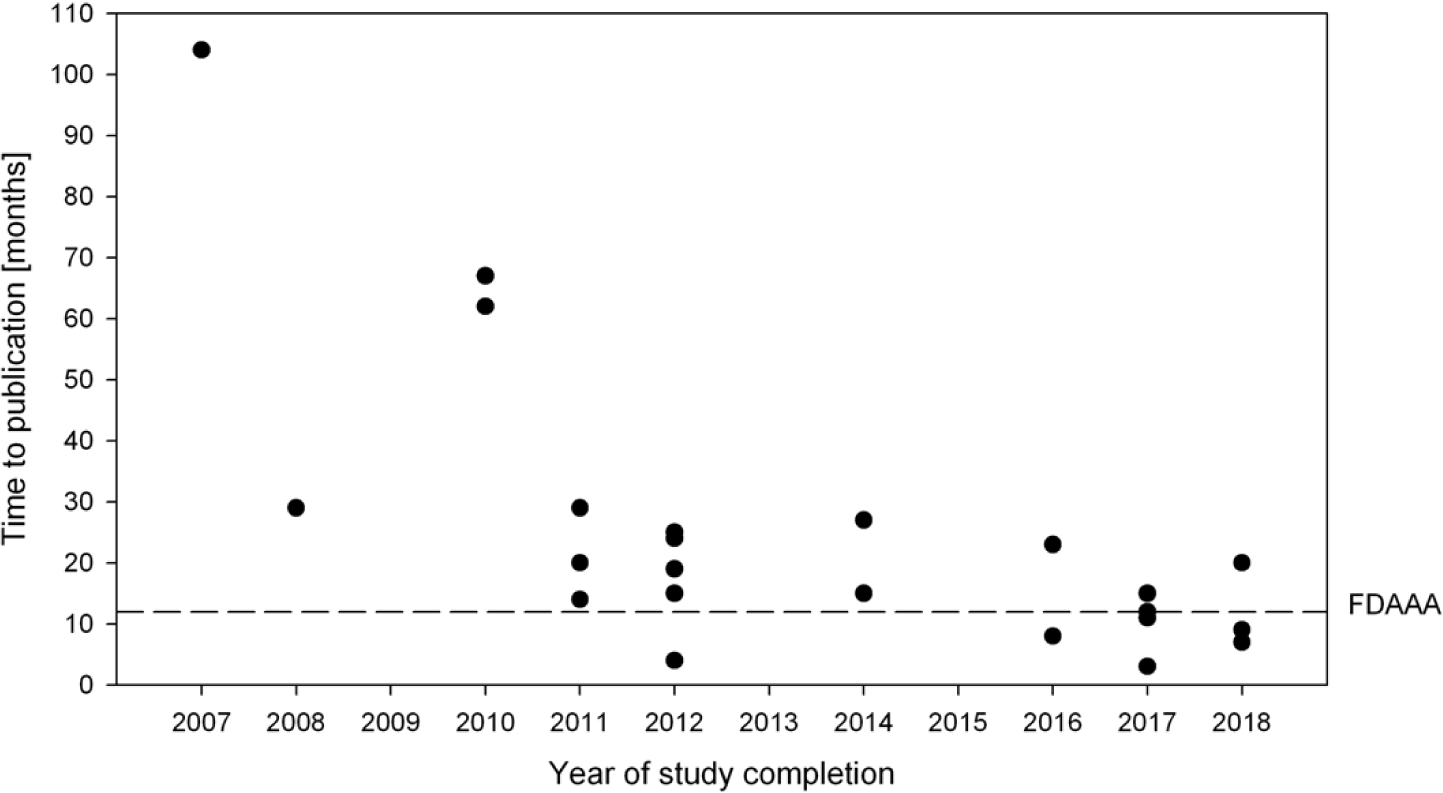
Time to publication (time between completion of the trial and publication of results) in months by year of completion “FDAAA” = timeline mandated by the U.S. Food and Drug Administration Amendments Act of 2007

## Discussion

Almost every second completed registered clinical trial on pediatric ED remains unpublished, making results from 1644 enrolled study patients unavailable for clinical decision making. Given the high prevalence of ED and its potentially serious Manifestations, this significant publication bias is both surprising and unsatisfying.

This lack of study results may directly influence clinical practice. An illustrative example is the use of dexmedetomidine. Two published studies could show a reduction of incidence and degree of ED following premedication with intranasal dexmedetomidine ^9 10^. However, dexmedetomidine, like most potent sedatives, causes an unpleasant burning sensation when applied intranasally ^11^. Oral application might therefore be a better choice for anxious children. One recent study showed that 1 µg per kg oral dexmedetomidine for premedication provided satisfactory sedation levels, but was not effective in preventing ED ^12^. On the other hand, we identified an unpublished registered trial (NCT03357718) that used 2 µg/kg, so it is not known whether oral dexmedetomidine at higher doses might be as effective as intranasal application. Another unpublished study (NCT03171740) compared premedication with intranasal dexmedetomidine to oral midazolam. Intra- or postoperative dexmedetomidine application was investigated in five registered trials (NCT01901588, NCT03779282, NCT00857727, NCT01895023, NCT01535287) the results of which are not available (yet) to the public. Especially with regard to different doses and potential cardiocirculatory side-effects of intravenous dexmedetomidine, the data of these 482 patients would be very interesting.

Similar considerations can be made for several study topics summarized in table 2. Minimizing pain with intraoperative Fentanyl given at a mean dose of 2.5 µg/kg at the end of surgery reduced the incidence of ED in a study by Cohen et al. ^13^. However, in the context of postoperative delirium in the PICU we could recently show that fentanyl increases the risk for delirium in a dose-dependent way and that this could probably attributed to substance-specific anticholinergic effects ^14^. Therefore it would be very interesting to see the results of the 322 patients from the three unpublished registered trials (NCT02753725, NCT03010540, NCT03062488) on intraoperative fentanyl given at different doses.

Unfortunately, the low publication rate for studies on ED that we found in our analysis is in line with other published observations. Anderson et al. recently reported that only 38.3% of all completed or prematurely terminated trials registered at ClinicalTrials.gov were published ^15^, and we came to similar conclusions when testing for publication bias in fields as diverse as pediatric liver transplantation ^16^ or autism ^17^. Publication of the results gathered in clinical trials involving human subjects is considered an ethical imperative ^18^. In 2007 it became a legal obligation in the U.S. to register all clinical trials in advance and publish its results within 12 months after completion ^8^. Interestingly, the highest rate of unpublished study with regard to the country of the investigation was found for the U.S. despite of this federal law. Timely publication of the results is another issue that we investigated in our study. Only 9 of the 24 published studies were published within 12 months after completion, and we did not observe a trend to shorter publication intervals during recent years.

### Limitations

Our study has several limitations. First, we only analyzed clinical trials that were registered either at ClinicalTrials.gov or ClinicalTrialsRegister.eu., therefore some studies registered in smaller national registers may have been missed. Second, our analysis relies on the accuracy of data input in the respective register. Third, we can only speculate about the reasons why half of the investigators chose not to publish their results, because we did not receive respective information after contacting them directly. Last, it is likely that some of the recently completed studies will be published eventually, but still considerably later than the 12 months warranted by the FDAAA.

### Conclusion

There is a distinct publication gap in clinical research in pediatric ED. Although this does not call into question the results of published studies, it should raise awareness that many aspects of the current treatment options are not exactly known and that larger numbers of published trials are immensely helpful to either support existing data or to challenge it thereby improving clinical practice. In addition, timely publication of study results helps to improve patient care and avoids unnecessary exposure to research if a similar research question in being investigated repeatedly.

## Data Availability

All relevant data are in the manuscript.

## Authors’ contributions

Substantial contributions to the conception or design of the work; or the acquisition, analysis, or interpretation of data for the work: JM, MR

Drafting the work or revising it critically for important intellectual content: JM, MR

Final approval of the version to be published: JM, MR

Agreement to be accountable for all aspects of the work in ensuring that questions related to the accuracy or integrity of any part of the work are appropriately investigated and resolved: JM, MR

## Data sharing statement

All relevant data are in the manuscript.

## Competing interests statement

JM and MR report no conflict of interest

## Notes

### Competing Interest Statement

The authors have declared no competing interest.

### Clinical Trial

There is no trial ID, because this study is not a clinical trial

